# Abnormal Intraepidermal Nerve Fiber Density in Disease: A Scoping Review

**DOI:** 10.1101/2023.02.08.23285644

**Authors:** SJ Thomas, J Enders, A Kaiser, L Rovenstine, L Heslop, W Hauser, A Chadwick, DE Wright

## Abstract

**Background:** Intraepidermal nerve fiber density (IENFD) has become an important biomarker for neuropathy diagnosis and research. The consequences of reduced IENFD can include sensory dysfunction, pain, and a significant decrease in quality of life. We examined the extent to which IENFD is being used as a tool in human and mouse models and compared the degree of fiber loss between diseases to gain a broader understanding of the existing data collected using this common technique.

**Methods:** We conducted a scoping review of publications that used IENFD as a biomarker in human and non-human research. PubMed was used to identify 1,004 initial articles that were then screened to select articles that met the criteria for inclusion. Criteria were chosen to standardize publications so they could be compared rigorously and included having a control group, measuring IENFD in a distal limb, and using protein gene product 9.5 (PGP9.5).

**Results:** We analyzed 397 articles and collected information related to publication year, the condition studied, and the percent IENFD loss. The analysis revealed that the use of IENFD as a tool has been increasing in both human and non-human research. We found that IENFD loss is prevalent in many diseases, and metabolic or diabetes-related diseases were the most studied conditions in humans and rodents. Our analysis identified 74 human diseases in which IENFD was affected, with 71 reporting IENFD loss and an overall average IENFD change of -47%. We identified 28 mouse and 21 rat conditions, with average IENFD changes of -31.6 % and - 34.7% respectively. Additionally, we present data describing sub-analyses of IENFD loss according to disease characteristics in diabetes and chemotherapy treatments in humans and rodents.

**Interpretation:** Reduced IENFD occurs in a surprising number of human disease conditions. Abnormal IENFD contributes to important complications, including poor cutaneous vascularization, sensory dysfunction, and pain. Our analysis informs future rodent studies so they may better mirror human diseases impacted by reduced IENFD, highlights the breadth of diseases impacted by IENFD loss, and urges exploration of common mechanisms that lead to substantial IENFD loss as a complication in disease.

## Introduction

The peripheral nervous system provides critical feedback and information from diverse somatic and visceral tissues, including muscles, joints, viscera, and the exterior surface. When damaged, peripheral nerves can degenerate and later regenerate to restore function to peripheral tissues [1]. Successful regeneration requires sophisticated communication between glial cells, vascular components, connective tissues, and the immune system [2, 3] and is dependent on energetic states [4-10]. New information is emerging related to the molecular signals required for what is now recognized as active processes leading to the degeneration of peripheral axons, including sterile alpha and TIR motif containing 1 (SARM1) and other potential mediators [11-14].

Additionally, recent advances have improved our understanding of specialized Schwann cells and their role in epidermal innervation, nerve retraction, and pain [15]. Overall, exciting progress is being made related to our understanding of the dynamic processes undergone by peripheral axons and potential therapeutic tools in neuropathy [16-19].

Epidermal innervation specifically has become a point of interest in several diseases, including diabetes and chemotherapy-induced neuropathy. The epidermis is richly innervated by unmyelinated C-fiber axons that cross the dermal-epidermal border and extend between keratinocytes to more superficial depths. Epidermal axons express numerous receptors and have specialized molecular profiles that allow them to respond to specific stimuli [20-22]. These axons degenerate in tissue injury and disease settings. However, our understanding of the normal plasticity of epidermal axons is premature. The longest sensory axons are often the first to undergo axonal loss, causing symptoms in the distal limbs in what is called a “length-dependent” neuropathy [23]. Loss of peripheral nerve axons in these areas is associated with decreased temperature sensation and pain, often described as burning or electric shock-like [24]. These epidermal axons do not function in isolation, as communication between keratinocytes, Schwann cells, and immune cells is likely essential in modulating their function in somatosensation and pain [25, 26].

The ability to visualize epidermal axons has added a vital avenue in understanding and diagnosing peripheral nerve degeneration. The emergence of PGP 9.5 as a tool that allows for consistent quantification of epidermal axons has dramatically accelerated this research [27, 28]. In addition, the relative ease and safety of obtaining a skin punch biopsy to elucidate IENFD has proven advantageous over more invasive biopsy approaches. Understanding the scope and importance of IENFD as a research tool is essential as we search for biomarkers that can inform disease progression and recovery.

In this scoping review, we assessed the current literature to determine the extent of IENFD use as an objective biomarker in human and non-human research and provide a quantitative overview of the severity of fiber loss found in different diseases. This review highlights the rapidly increasing use of IENFD measurements in clinical and basic research within the last 20 years and the diverse array of diseases in which IENFD has been measured. We also examine the similarities and differences that have emerged between human and non-human research concerning IENFD measurement. Finally, we compare changes in IENF densities in subtypes of diseases such as diabetes and chemotherapy to provide additional insight into how these disease subtypes differentially affect epidermal axons.

## Methods

### 1. Research question

This review aimed to determine the scope of animal and human conditions in which experimental studies have used IENFD to measure and compare percent fiber change. It was conducted in the following five key phases: 1) outlining the research aim, 2) identification of records matching search terms, 3) initial screening for relevant articles, 4) in-depth screening and data collection, 4) visually mapping the data, 5) describing, summarizing, and reporting the results.

### 2. Literature collection and screening

Our literature screening and collection process is outlined in **Fig 1**. The initial search was conducted in July of 2022 using PubMed as the single data source. The search term consisted of the following string: “intraepidermal nerve fiber density OR epidermal nerve fiber density OR IENFD OR IENF NOT (Review[Publication Type]) NOT (Case Reports[Publication Type]), NOT (Systematic Review[Publication Type]) NOT (Meta-Analysis[Publication Type])”. The abstracts of the remaining 1,004 records were manually screened for general relevance, including measuring IENFD, conducting original research, and written in English.

**Figure 1:**
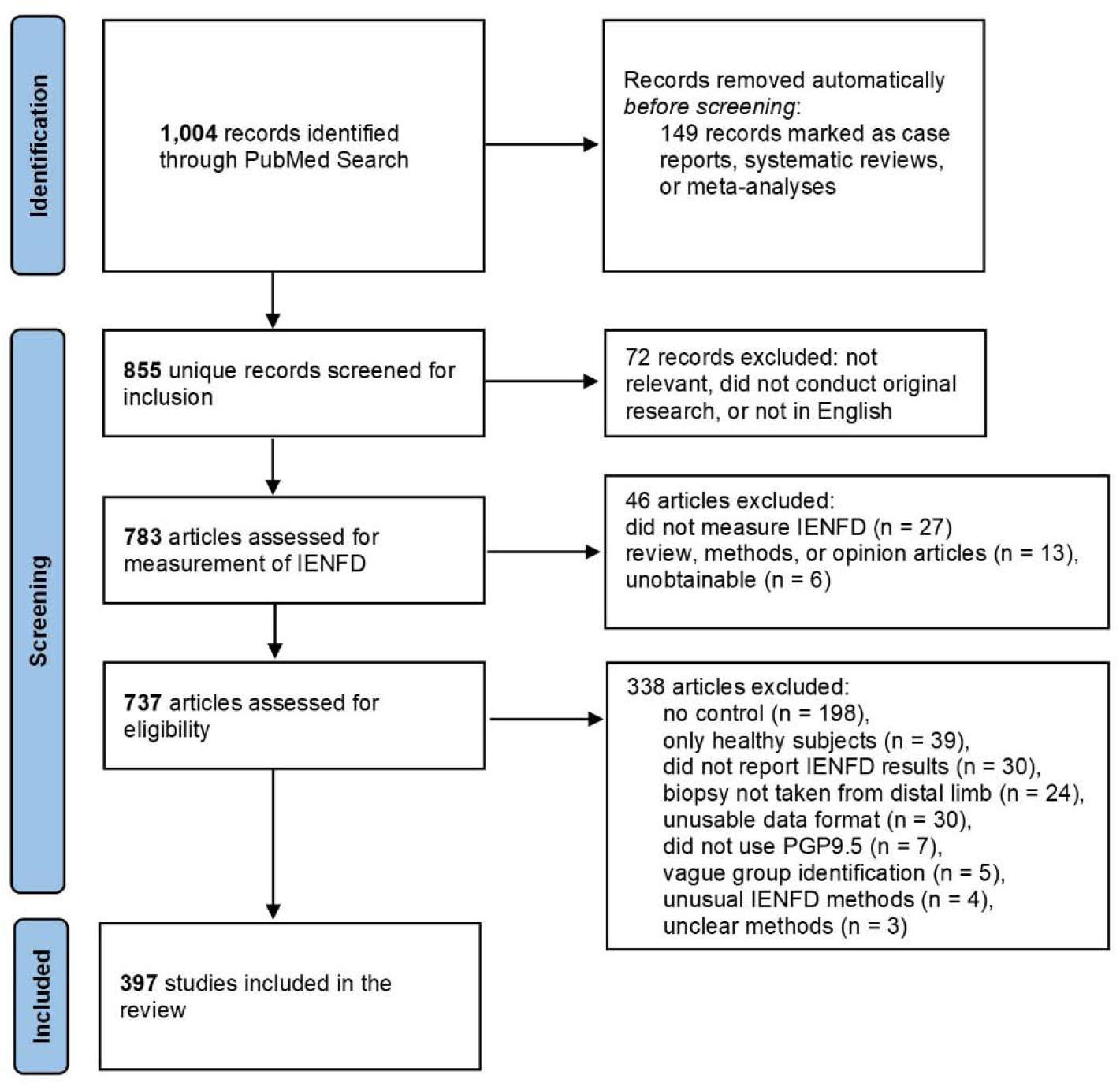
PRISMA flowchart of the article identification, screening, and inclusion process.

After this initial screen, all articles were downloaded and assessed to see if they fulfilled the following inclusion criteria: having an experimental group and control cohort for comparison, measuring IENFD at a distal limb, and staining axons using PGP9.5. These criteria were selected to provide consistency among the collected data and rigorously compare publications. Articles were excluded if the methods or experimental groups were unclear or if they used unusual methods for measuring IENFD such that they could not be compared to other articles. Publications that measured IENFD but did not fulfill these criteria were recorded and included in **Fig. 2** but were not included in the subsequent data analysis. All citations were managed using EndNote.

**Figure 2.**
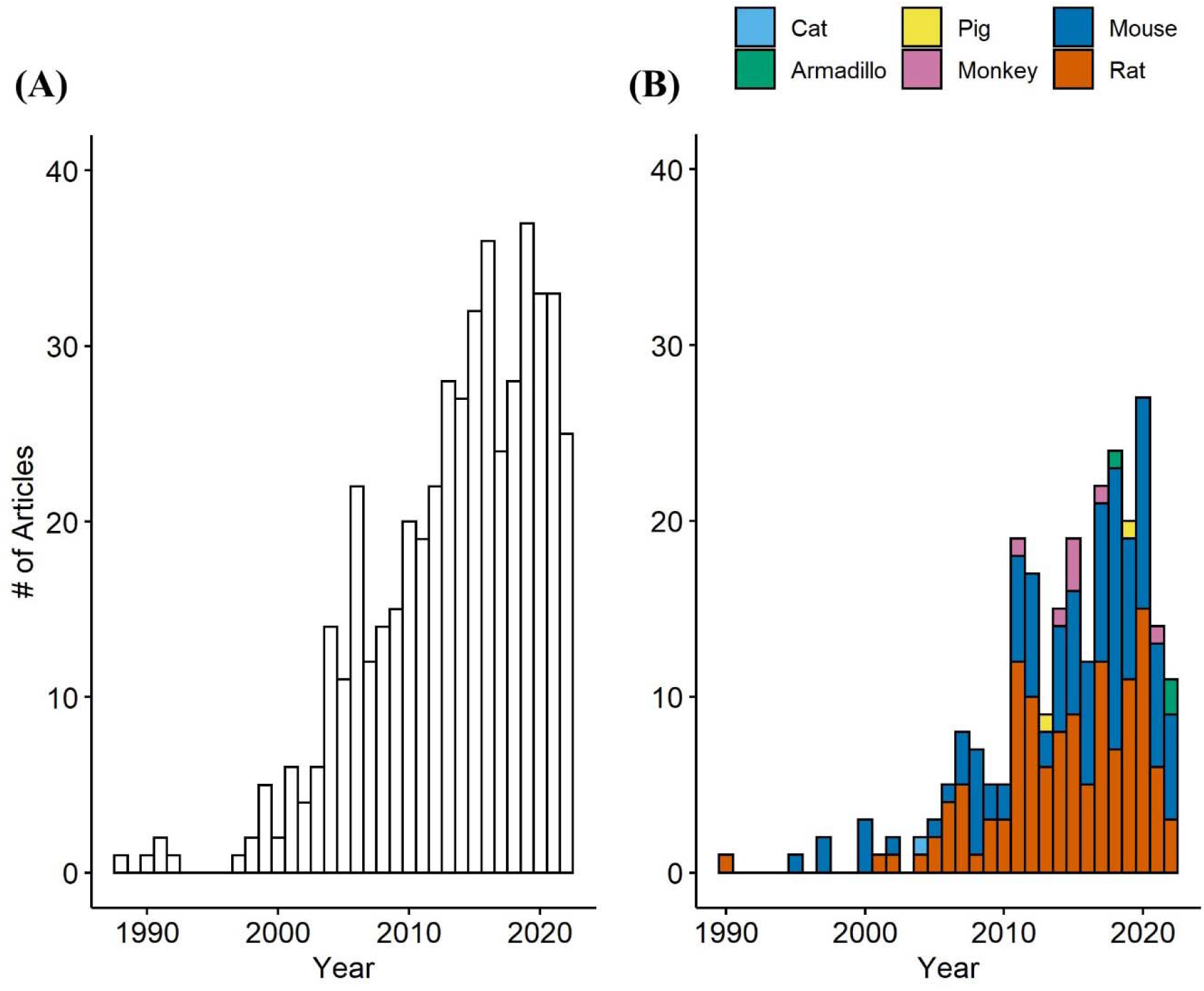
Number of papers measuring IENFD per year in humans (A) and non-human species (B). The number of papers measuring IENFD in non-human species is presented cumulatively for each year.

### 3. Data Collection

The percent IENFD loss was extracted from publications using two methods. The preferred method was to record the experimental and control group fiber density, or percent fiber loss, as reported directly by the publication. If values were not provided, ImageJ was used to measure the y-axis to the last tick mark and bar heights in pixels in cases where these values were reported graphically. The value assigned to the last tick mark of the y-axis was recorded and used to convert the pixel length of bars to y-axis units. If the y-axis did not start at zero, the paper was excluded. The recorded control and experimental group values were then used to calculate the relative percent IENFD change. For animal studies, if there were several time points in the study design, these were recorded separately and then averaged. Snapshots of the measured graphs and pixel values were captured and saved with each publication’s file in EndNote.

In addition to IENFD percent change, other data collected from articles included the year published, species and strain used, experimental and control groups, size of the experimental group if available, and biopsy location. All collected numerical data and recorded experimental groups were confirmed by a separate reviewer to check for accuracy.

### 3. Data Summary and Synthesis

The data collected from multiple reviewers were compiled into a single Microsoft Excel spreadsheet. The data was checked for consistency and implausible values. It was then exported into RStudio 4.2 to create graphical representations of the data and calculate percentages and frequencies. All data and code are available upon request.

## Results

### 1. Number of Studies Reporting IENFD Per Year

Our first goal was to determine how the frequency of IENFD use as a tool in research has changed over the last several decades. We analyzed the data set for the number of publications per year in both human and non-human species. Our search revealed publications as early as 1988 in human studies and 1990 in rats that utilized IENFD according to our criteria. Over the next ten years, intermittent publications emerged on humans and rodents. Starting around 2000, the number of publications reporting IENFD results steadily increased. Publications on human tissue (**Fig. 2A**) peaked at 37 papers in 2019, while non-human publications (**Fig. 2B**) peaked at 27 in 2020. This steady and rapid increase in publications reflects the increasing utility of IENFD in clinical and translational research.

Publications assessing IENFD in rodents dominate the non-human literature, with rats and mice comprising 49.6% and 45.3% of published non-human studies, respectively. Publications in non-rodent animals emerged later and now include armadillos, monkeys, pigs, and cats. Overall, these findings suggest that the recognition of IENFD’s value in animal research and the ease of IENFD measurement has increased over the past two decades.

### 2. Number of Unique Conditions Measured Per Year

The number of conditions in which IENFD has been measured has increased substantially over the past 30 years. In clinical research (**Fig. 3A**), 20 unique conditions were measured at their peak in 2020. Although the number of conditions in which IENFD was measured is higher in humans than in rodents, the diverse conditions in which IENFD has been measured in rodent species has also increased (**Fig. 3B**). This increase has been more pronounced in mice than rats in recent years. Mouse unique conditions peaked in 2018 with 11 conditions, while rats peaked in 2015 with seven conditions. This broadening interest indicates that IENFD is becoming relevant in increasingly diverse contexts.

**Figure 3.**
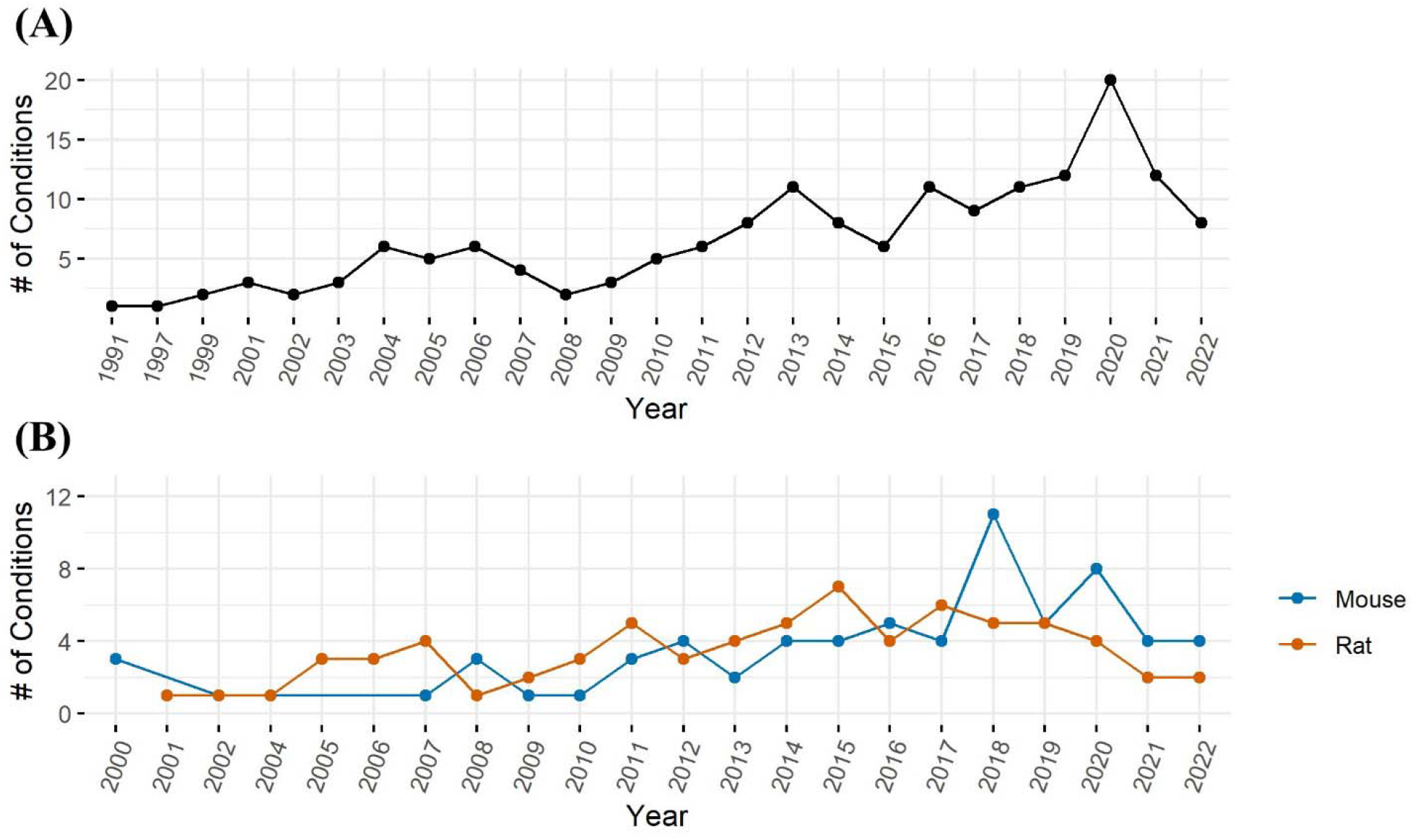
Number of unique conditions with IENFD measurements per year in humans (A) and rodents (B).

### 3. Commonly Studied Conditions and Disease Types

One important goal of the present review was to identify conditions where IENFD was commonly measured and organize them by disease category. IENFD changes in human subjects with diabetes mellitus were assessed in 107 publications, far more than for any other condition (**Fig. 4A**). Often, articles would not specify the type of diabetes mellitus, so distinguishing between type 1 and 2 in all cases was not possible. Several papers reported the condition they measured as simply “neuropathy”, either from multiple conditions or without further specifications of the cause. This general classification was the second most measured condition. Parkinson’s disease, chemotherapy, and other diseases also had IENFD measurements, albeit at a much lower frequency than in diabetes or neuropathy. Conditions were further categorized into one of six groups (**Fig. 4D**). The most common disease groups with IENFD measurements in humans were “metabolic” (37%), followed by “neurodegenerative” (25%), and “other” (18%).

**Figure 4.**
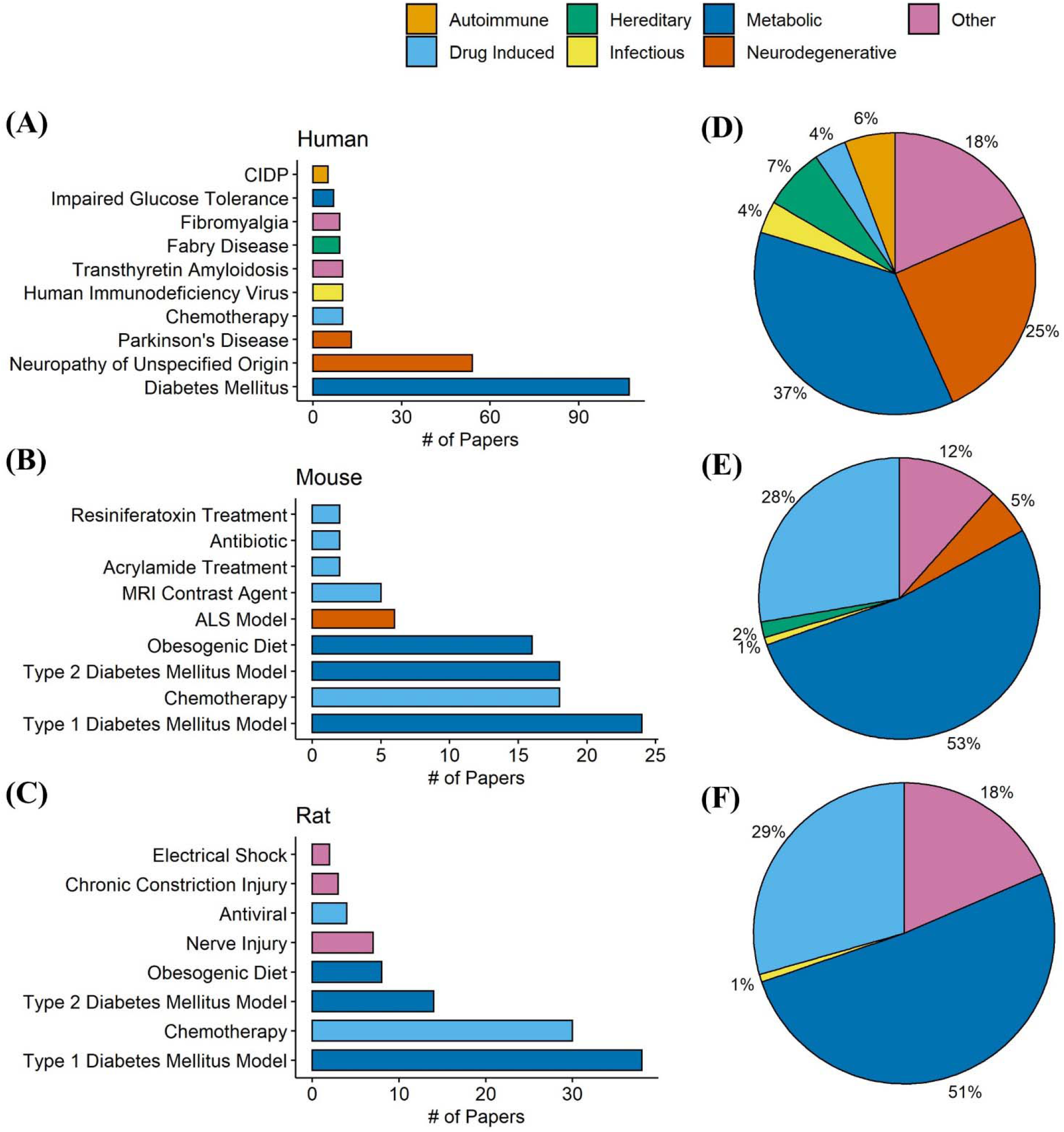
Total number of papers in our database measuring commonly studied conditions in humans (A), mice (B), and rats (C). Each condition is color coded by its designated condition category. Each condition was broadly classified into the categories autoimmune, drug induced, hereditary, infectious, metabolic, neurodegenerative, or other. D, E, and F represent the percentage of papers in our database that measured conditions in each respective category in humans, mice, and rats. The condition category classifications can be found in **Supplementary Table 1** for humans and **Supplementary Table 2** for rodents.

Similar to human studies, type I diabetes was the most frequently studied model in mice and rats, followed by chemotherapy, type II diabetes, and an obesogenic diet (**Fig. 4 B-C**). In contrast to human studies, over half of the conditions measured in mice (53%) and rats (51%) fell into the metabolic grouping (Fig. 4 **E-F**). Compared to publications in humans, there was a higher percentage of studies in mice or rats that investigated drug-induced conditions, such as chemotherapy and anti-microbial models. This analysis indicates that certain diseases, such as diabetes, are similarly measured in human and rodent models and provide a strong foundation for translational relevance. However, the frequency of disease categories examined using IENFD is not perfectly aligned between human and rodent studies.

### 4. IENFD Change in Human Diseases

We sought to compare percent fiber loss across human diseases captured in our data set. Seventy-four human conditions were identified in our data search, and axonal fiber loss is present at varying levels in 71 diseases or conditions (**Fig. 5**). The average IENFD change across all human conditions was -47%. The three diseases with the highest percentage of fiber loss were familial dysautonomia (−98.6%), leprosy (−95.8%), and amyloid neuropathy (−88.1%). Of all the diseases/states identified, 32% had greater than 50% fiber loss, and 74% had greater than 25% fiber loss. Interestingly, three diseases were identified that reported increases in IENFD. These diseases were Miller-Fisher syndrome (1.8%), psoriasis (31.9%), and Rett syndrome (51.6%), although psoriasis had wide standard deviation bars due to varying results from different studies. Although axonal fiber loss is classically associated with conditions like diabetes and chemotherapy-induced peripheral neuropathy, this data suggests that reduced IENFD loss occurs in many human pathologies.

**Figure 5.**
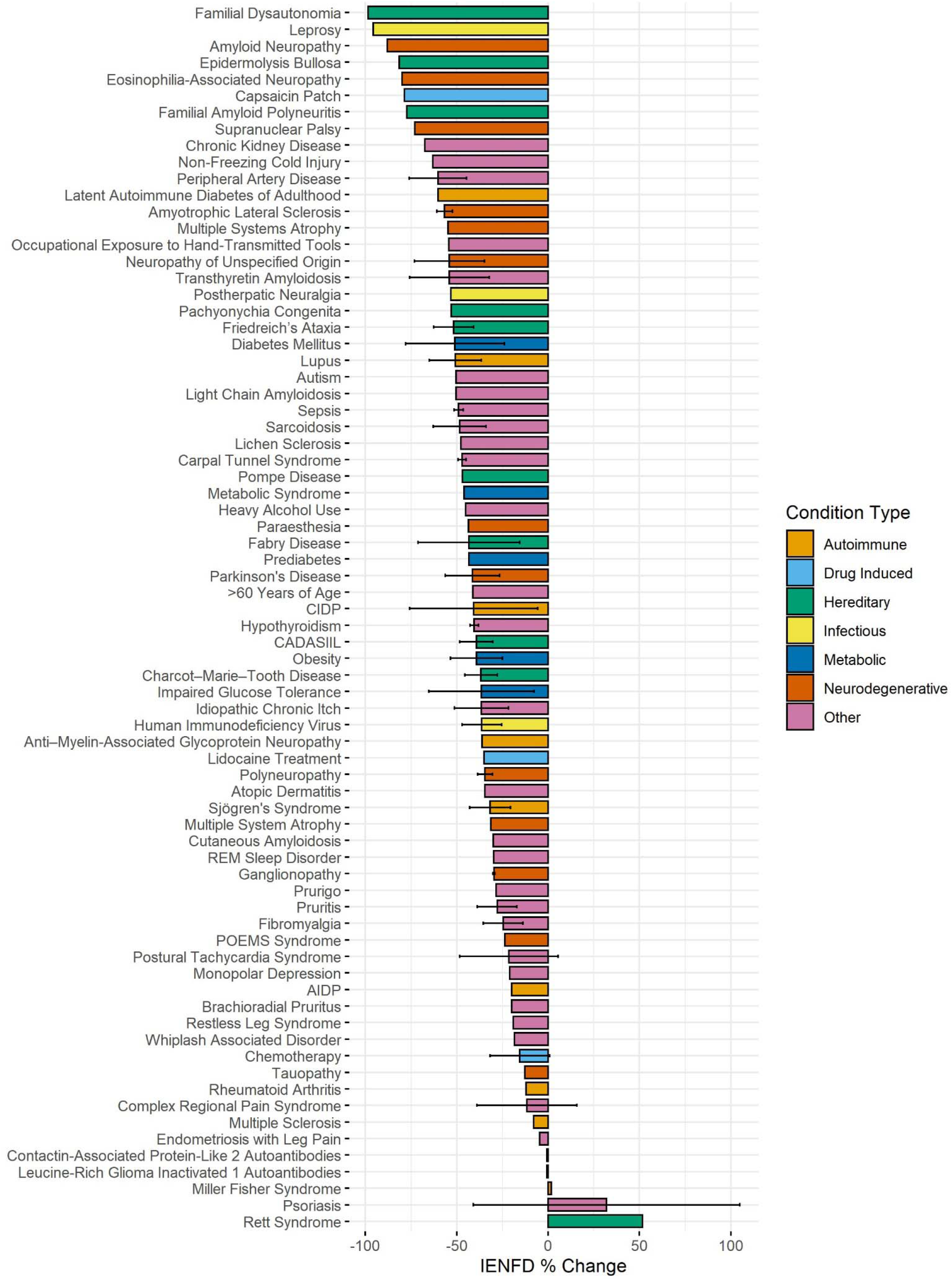
Average IENFD percent change relative to control for human conditions in our dataset, weighted by experimental group size with standard deviation bars. Each condition is color-coded by its condition category. CADASIL = Cerebral Autosomal Dominant Arteriopathy with Sub-cortical Infarcts and Leukoencephalopathy, POEMS = Polyneuropathy, Organomegaly, Endocrinopathy, Monoclonal protein, Skin changes.

### 5. Human Diabetes Mellitus Subtypes

Next, we used our data set to make comparisons within specific conditions. One hundred and seven papers examined diabetes in humans, allowing us to stratify based on different characteristics in each study. Our analysis found that type 1 diabetes had slightly higher levels of fiber loss than type 2 diabetes (−47.1% vs. - 42.2%, **Fig. 6A**). A more dramatic difference was present if neuropathy was reported. Diabetic patients with neuropathy had more than double the fiber loss of diabetic patients without neuropathy (−61.1% vs. -25.9%). Diabetic patients with kidney or pancreas transplants had the highest levels of neuropathy at -79.3%.

**Figure 6.**
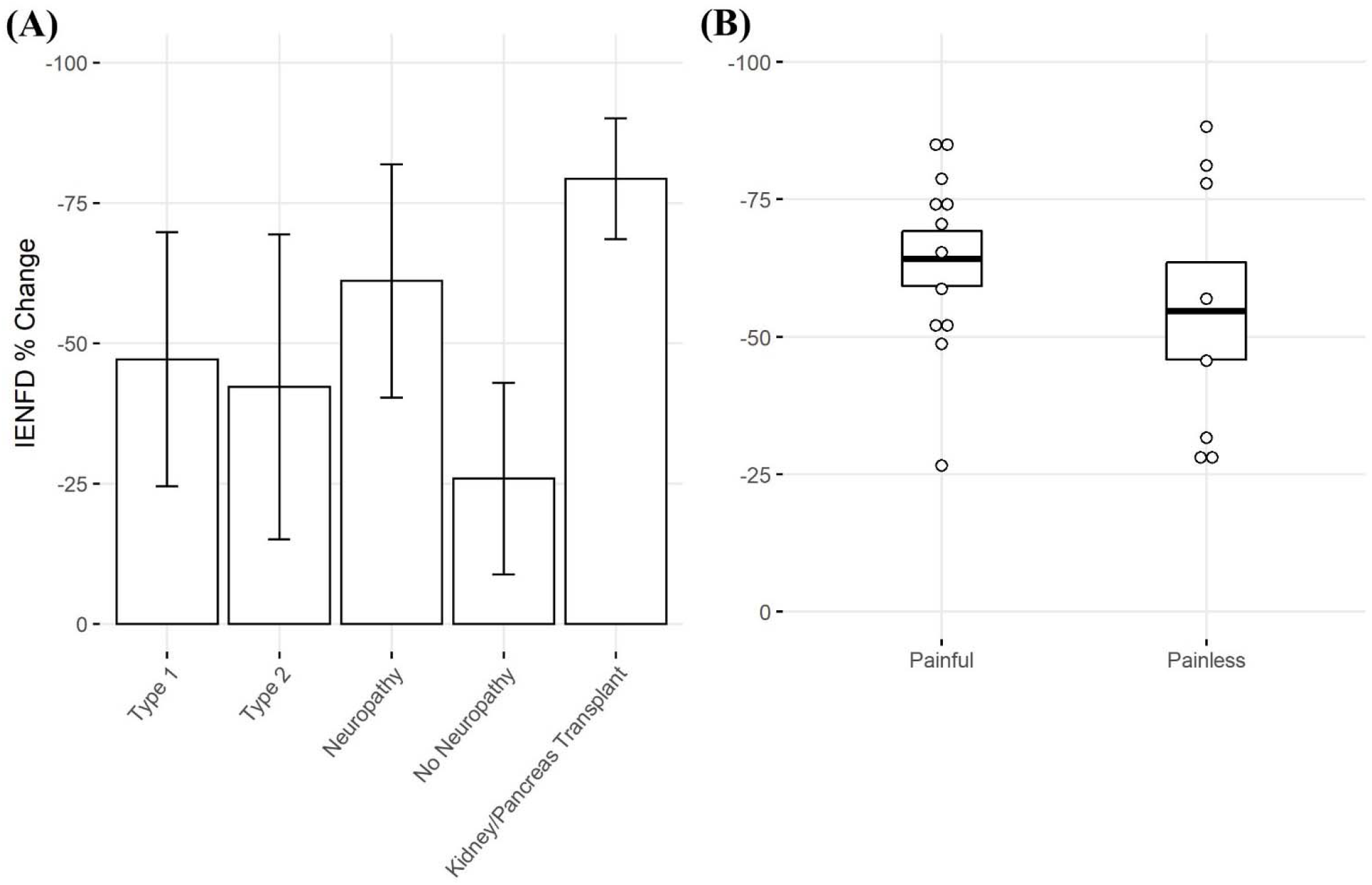
(A) Percent IENFD change compared to control in human subjects with diabetes mellitus by subtype, weighted by experimental group size with standard deviation bars. (B) Percent IENFD change in diabetes mellitus in painful vs painless subtypes with standard error bars and each data point representing an individual paper.

Within the subset of publications that specified whether diabetic neuropathy was painful or painless, those with painful neuropathy tended to have slightly higher levels of fiber loss (**Fig. 6B**). Painful and painless diabetic neuropathy had an average fiber loss of -64.2% and -54.7%, respectively.

### 6. Chemotherapeutic Agents in Humans

We also examined fiber loss due to different chemotherapy treatments in humans (**Fig. 7**). A wide range of fiber loss can be seen, likely due to varying time points and doses in the studies. A single study reported reduced IENFD following treatment with Taxol (−27.6%). Oxaliplatin was the most studied human chemotherapy drug and had an average loss of -19.9%. Finally, bortezomib had the lowest average fiber loss at -8.9%.

**Figure 7:**
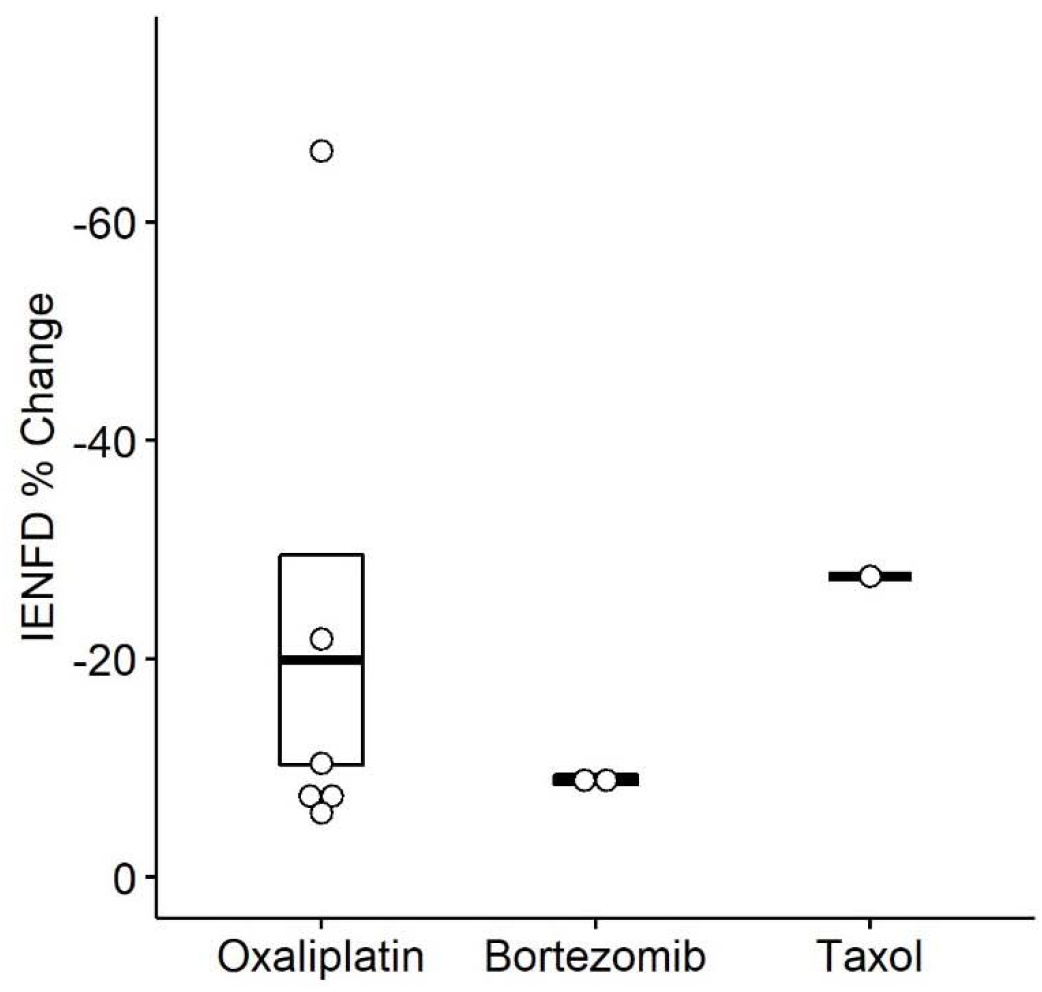
Percent IENFD change compared to control in humans in response to different chemotherapy treatments with standard error bars and each point representing an individual paper.

### 7. IENFD Change in Rodent Conditions

Rodent studies also showed axonal fiber loss in several diseases and conditions. In our data set, we identified 28 different conditions in mice (**Fig. 8A**) and 21 in rats (**Fig.8B**) in which IENFD was reported. The average IENFD change was -31.6 % and -34.7% among mouse and rat conditions, respectively. Nerve injury (− 98.2%), femoral artery ligation (−71.9%), and chronic constriction injury (−70.9%) caused the highest severity of fiber loss in mice. Rat models did not reach the same overall severity of fiber loss, with 100g weight traction injury (−65.7%), electrical shock (−61.6%), and varicella-zoster virus infection (−60.7%) as the three most deleterious conditions. In both species, multiple conditions caused an increase in fiber density. These experimental approaches in mice included a ketogenic diet (5.7%), hypoxia-inducible factor knock-out (6.1%), insulin receptor knock-out (9.1%), dry skin (85.2%), and atopic dermatitis (129.8%) (**Fig. 8A**). In rats, these included antibiotic treatment (specifically minocycline, 7.4%), complement Freund’s adjuvant (70%), dermatitis (74.3%), and dry skin (111.4%) (**Fig. 8B**).

**Figure 8.**
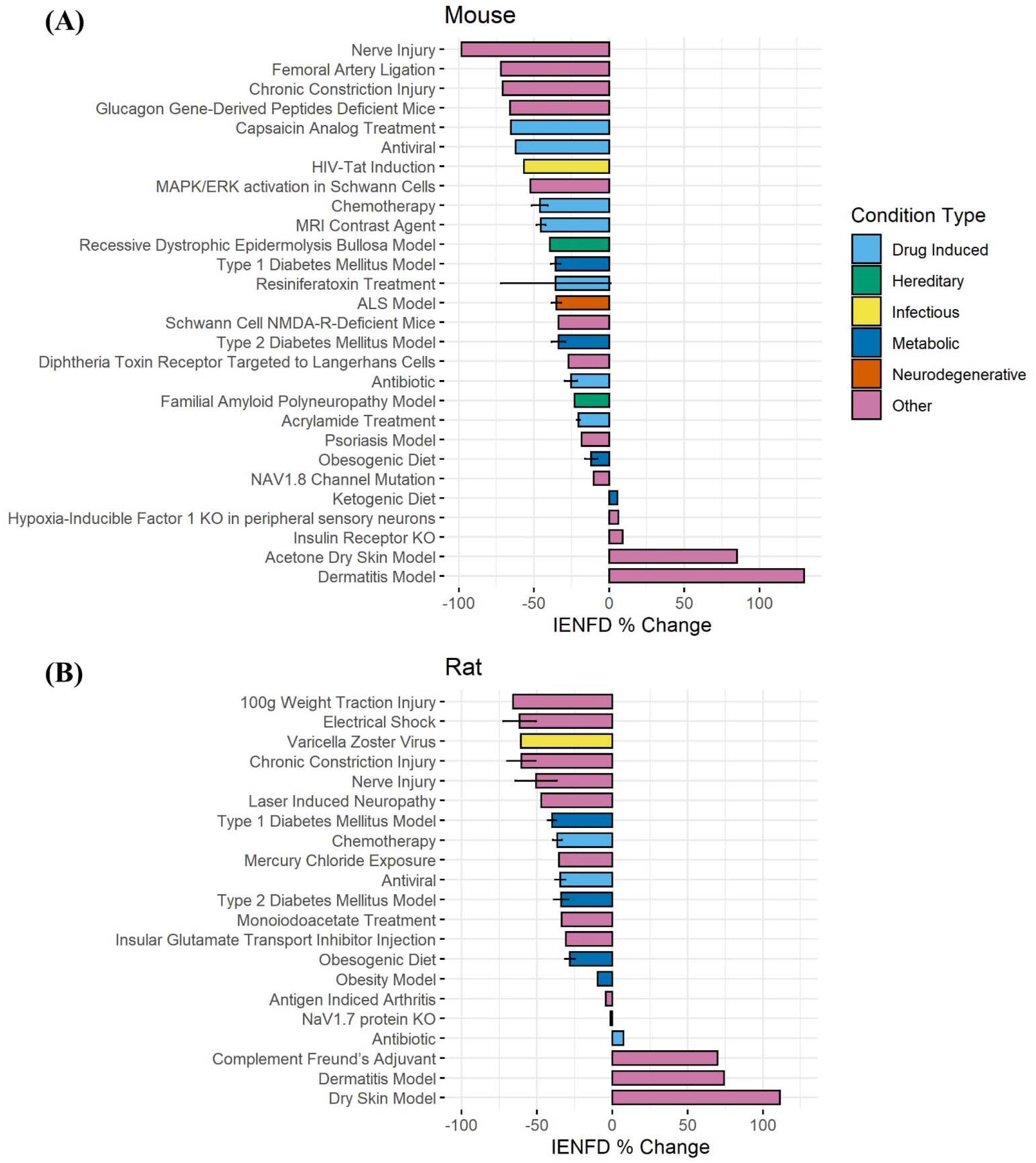
Average IENFD percent difference from control for each condition in mice (A) and rats (B) in our dataset with standard error bars. Each condition is color-coded by its condition category. ALS = amyotrophic lateral sclerosis, KO = knock out.

### 8. Rodent Diabetes Mellitus Subtypes and Models

Four main animal models of diabetes were identified and examined for their differences in fiber loss in mice and rats (**Fig. 9A**). In mice, STZ injection plus a high-fat diet resulted in the most significant reduction in IENFD (−36.1%), while a high-fat diet alone produced the least IENFD loss (−16.3%). In rats, STZ alone narrowly produced the greatest IENFD loss (−39.9%), followed closely by STZ injection plus a high-fat diet (−38.9%). A high-fat diet produced the least IENFD loss (−30%). The percent fiber loss due to STZ and STZ plus high-fat diet was similar in both mice and rats; however, a high-fat diet alone produced more significant IENFD loss in rats than in mice. STZ treatment alone had the most variable IENFD loss across all rodent studies.

**Figure 9.**
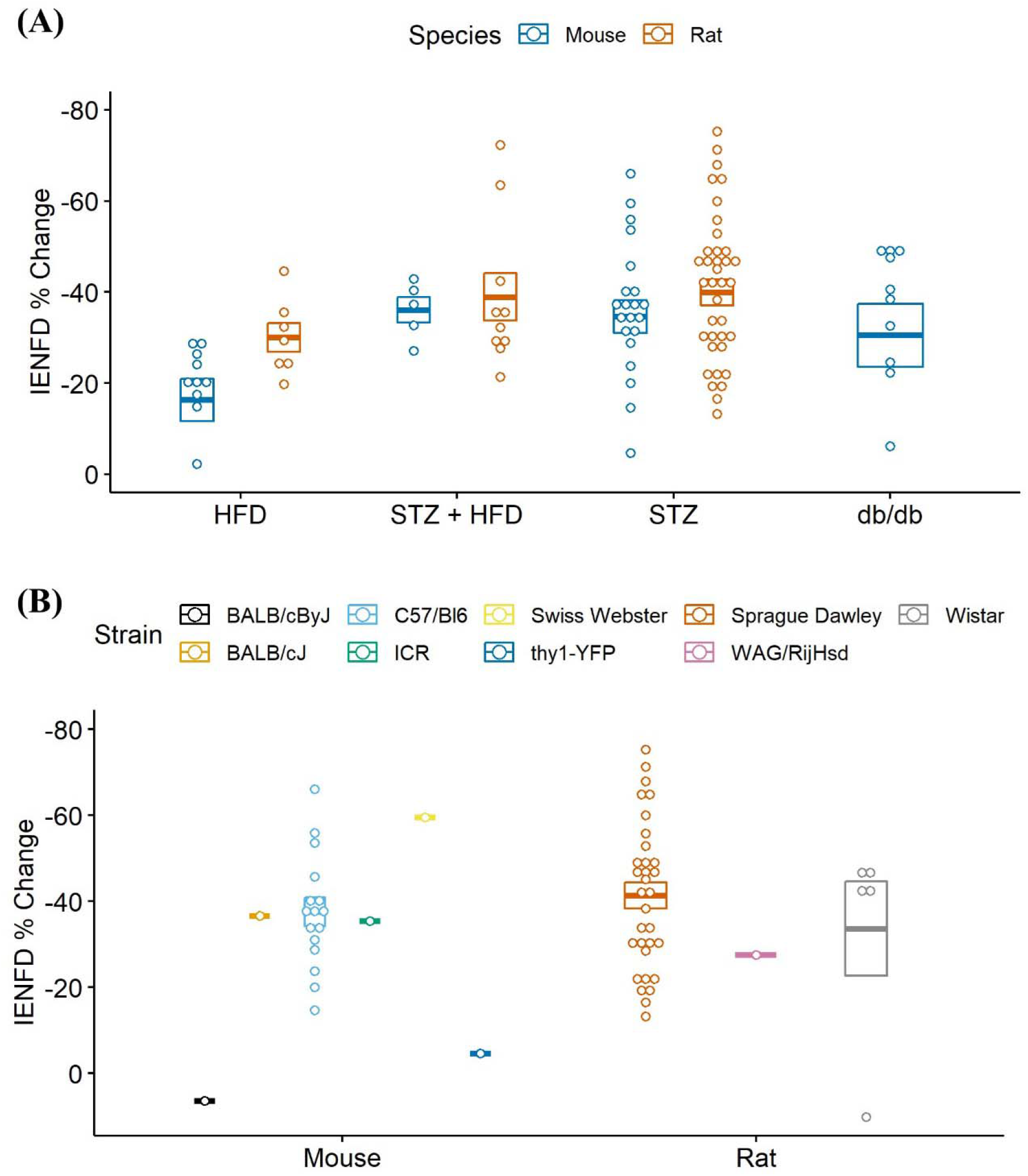
(A) Percent IENFD change from control in different models of diabetes in mice and rats with standard error bars and each point representing an individual paper. (B) Percent IENFD change in STZ induced diabetes, a common type 1 diabetes model, furthur broken down by strain of mouse or rat used in each study. HFD = high fat diet, STZ = streptozocin.

When analyzed by rodent strain, STZ treatment alone did not produce a clear picture of which rodent strains are most vulnerable to IENFD loss (**Fig. 9B**). The most commonly studied strain was C57/Bl6 in mice and Sprague Dawley in rats. Our analysis suggests that BALB/cByJ (6.4%) and thy1-YFP (−4.6%) are protected from STZ-induced IENFD loss, but this was only demonstrated in one study for each model. A single study using Swiss Webster mice had the highest average IENFD loss due to STZ treatment (−59.5%). Sprague Dawley (−41.3%), WAG/RijHsd (−27.5%), and Wistar (−33.6%) rats tended to have similar IENFD loss, but one data point in Wistar rats was dramatically lower than the others.

### 9. Chemotherapeutic Agents in Rodents

We also compared IENF loss in various chemotherapy treatments in rodents (**Fig. 10**). Rats experience the highest average IENFD loss in response to oxaliplatin (−46.4%) and the least in response to cisplatin (−29.7%). Mice had the highest percent IENFD loss in response to Taxol (−61.8%) and, like rats, had the least in response to cisplatin (−27.9%). Mice, on average, had more severe IENFD loss in response to oxaliplatin and Taxol than rats. Cisplatin appeared to have a similar effect on the percent IENFD loss in both mice and rats.

**Figure 10.**
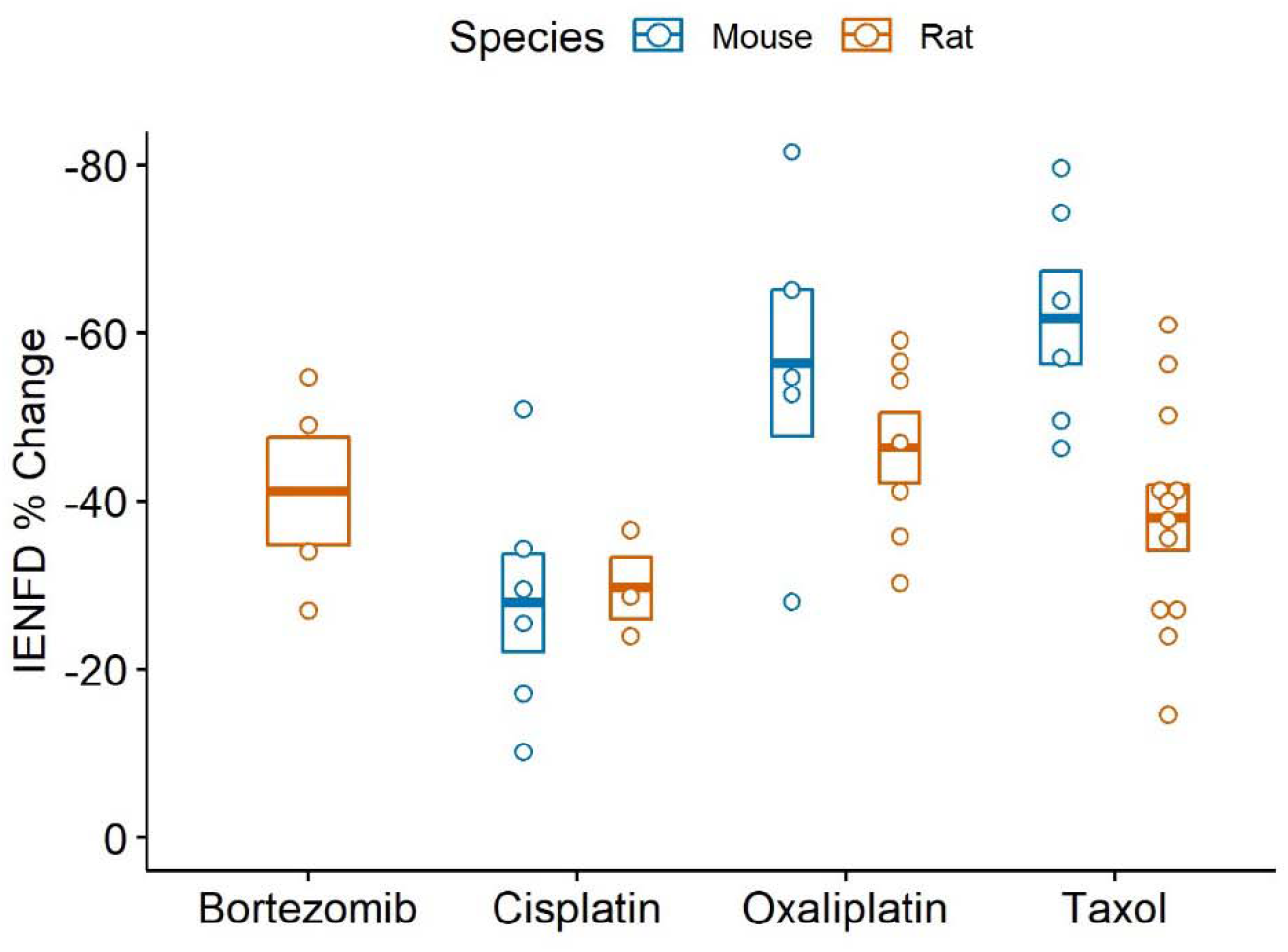
Percent IENFD change relative to control in different chemotherapy treatments in rodents with standard error bars and each point representing an individual study.

## Discussion

We performed a scoping review of the current literature to assess how IENFD quantification is used as a research tool and compare the amount of fiber loss across various conditions. One takeaway from this analysis is the rapid increase in the utilization of IENFD as a diagnostic tool in research. IENFD use increased by 65% in human studies and 440% in non-human studies from 2010 to 2020. IENFD assessments peaked around 2020 in both human and non-human studies, indicating that IENFD is an increasingly valuable tool in the field that may continue to gain widespread use in the future. Additionally, human studies continue to take advantage of IENFD measurements more frequently than non-human studies. This may be because IENFD quantification was first standardized in human skin biopsies [29, 30]. Better standardization in non-human IENFD measurements and reporting will improve generalizability and is critically important for translational relevance to non-human studies.

Another important finding of this analysis is the appreciation of the breadth of diseases with comorbid IENFD loss. Within the constraints of our analysis, we identified 74 human conditions in which IENFD was quantified in a standardized manner using PGP 9.5. Reduced IENFD loss was a feature of 71 publications (96%). The number of novel diseases or conditions measuring IENFD has been increasing similarly in both human and rodent publications (**Fig. 3**). Although the number of conditions measured in rodents has increased, our review identified approximately 50% more conditions with IENFD assessment in humans than in rodents. The types of diseases being measured in rodents also do not closely reflect those of humans. Metabolic disorders make up over 50% of rodent research but only represent 37% of human conditions with IENFD assessment (**Fig. 4**). Additionally, neurodegenerative disorders make up 25% of conditions studied in humans but comprise less than 5% of rodent studies, none of which occurred in rats. A small number of human studies examine IENFD loss resulting from autoimmune disease (6%); however, none were present in rodent studies. These differences highlight a potential disconnect between human and animal research and the complexity of modeling human disease in rodents.

Unsurprisingly, diabetic patients with neuropathy experienced greater IENFD loss than those without neuropathy in humans. This is expected, as sensory axon damage is intrinsic to peripheral neuropathies and leads to increased severity and incidence of small fiber neuropathy [1]. Diabetes patients who received a kidney or pancreas transplant tended to have the greatest IENFD loss, likely reflecting the severity of diabetes and possibly identifying additional toxins that damage epidermal axons. Although painful diabetic neuropathy had only modestly more IENFD loss than diabetic neuropathy without pain, the severity of fiber loss in painless neuropathy exhibited greater variability across studies (**Fig. 6B**). Thus, greater fiber loss may unreliably correlate with pain in diabetes. This remains an essential question in diabetes-related pain, as it is currently unclear why some patients with diabetes are more prone to developing pain than others.

The sub-analysis of different chemotherapeutic agents in humans yielded variable results, making the data difficult to interpret. This was likely due to the smaller number of studies and variability in drug dose and timing. Further research comparing the extent of fiber loss caused by different chemotherapeutic agents in humans may provide insight into common or diverse mechanisms.

The sub-analysis of rodent models of diabetes revealed that both STZ injection, a type 1 model, and low-dose STZ plus a high-fat diet, a type 2 model, produced approximately a 40% IENFD loss in both mice and rats. Average fiber loss for *db/db* mice, a diabetic model caused by leptin receptor deficiency, exhibited slightly lower but comparable levels of IENFD loss. The severity of IENFD loss in rodents is similar to that of type 1 and 2 diabetes in humans, supporting the translational power of these models related to axon degeneration. Consumption of a high-fat diet in mice did not lead to as severe IENFD loss as in other metabolic models. Administration of a high-fat diet caused about a 10% greater fiber loss in rats than in mice, producing IENFD loss levels closer to that of the other experimental models. This may indicate that rats are more susceptible to IENFD loss in a high-fat diet model than mice.

Our analysis confirmed that numerous metabolic diseases and conditions affecting mitochondrial and energetic health have IENFD loss as a complication. However, this review also illustrates how broadly applicable this approach can be in identifying other systemic diseases that may have common comorbid mechanisms that lead to axon degeneration. For example, this work identified various neurologic conditions, including diseases that feature central nervous system degeneration. It remains unclear how central nervous system degeneration affects peripheral sensory innervation. Potential mechanisms include a “top-down” effect driven by central nervous system degeneration. For example, it has been found that direct injection of glutamate into rat insula with no other intervention led to IENFD reductions, highlighting the possibility that imbalances in CNS excitatory neurotransmitters may play a role in IENFD [31]. Alternatively, a “top-around” effect in which signaling molecules or factors are released due to CNS injury could secondarily impact peripheral fibers through the bloodstream or other nonneural pathways [32-36].

The conditions identified by this review also suggest that the maintenance of peripheral fibers in the skin is critically sensitive to infectious or autoimmune threats. Loss of IENFs is largely dependent on Wallerian degeneration and SARM1 activity [37-40]. While direct links between inflammation and SARM1 activity in the peripheral nerve are emerging [41, 42], SARM1 is known to be heavily involved in responses evoked by pathogen- and damage-associated molecular patterns (DAMPs and PAMPs) [43, 44]. Combined with the breadth of infectious and autoimmune diseases exhibiting IENF loss described here, this suggests interplay between pattern recognition receptors and the Wallerian degeneration pathway may contribute to axon retraction. As future studies explore this possibility, a broader range of infectious or autoimmune conditions with accompanying fiber loss and small fiber-associated symptoms, including pain, will likely emerge.

Another well-represented area involving IENFD loss included conditions with hereditary genetic abnormalities. It will be important to align these hereditary conditions with mechanisms that lead to axon degeneration, as these mechanisms would likely be conserved across conditions featuring IENFD loss. For example, subtypes of Charcot-Marie-Tooth disease (CMT) include impaired axonal transport and mitochondrial function [45-48]. Notably, these consequences are implicated in chemotherapy-induced peripheral neuropathies [49-52] and neuropathies associated with diabetes and metabolic syndrome [53-57]. Impaired axonal transport is more amenable to the traditional model of Wallerian degeneration, in which disruption of distal nicotinamide-adenine dinucleotide (NAD^+^) homeostasis and energetic deficiencies lead to SARM1 activation [17, 58-60].

The extent to which these mechanisms converge in CMT, chemotherapy-induced peripheral neuropathy, and DPN is unclear and should be explored. The similarity across these etiologies is notable and suggests that conserved mechanisms underlying small fiber pathologies may be more common than initially thought. This scoping review helps to shed light on links between degenerative mechanisms that include comorbid IENFD loss across a broad range of diseases and conditions.

Overall, measuring epidermal fiber density is a relatively simple and non-invasive approach that can provide a window into the health of sensory axons in the epidermis. This analysis helped us understand the breadth of IENFD loss in a diverse array of human diseases and provided insight into how IENDF is being used as a translational tool in non-human research models. An important limitation of this analysis is that the disease duration in human and non-human experimental research was not included. Increased disease duration impacts axon loss, and much of the variability between similar studies was likely due to differences in the disease duration or stage at which IENFD was measured. Another limitation of our analysis is that drug dosage was not considered in studies that utilized pharmaceuticals. The same drug may have different effects on IENFD at different doses, and this review could not capture those distinctions. Finally, our literature search likely did not find every paper that performed IENFD measurements. It is possible that we only captured a sample of the research that has been performed in this area, and our data may not display the complete picture of the current literature. However, our analysis likely captures a representative snapshot of the field and provides a useful overview of IENFD in research and the severity of IENFD loss across different diseases, non-human models, species, and drugs.

## Data Availability

All data produced in the present study are available upon reasonable request to the authors.

## Conflict of Interest

The authors declare that the research was conducted in the absence of any commercial or financial relationships that could be construed as a potential conflict of interest.

## Funding

This work was supported by NIH grants RO1 NS043314 (DEW), the Kansas Institutional Development Award (IDeA) P20 GM103418, Kansas University Training Program in Neurological and Rehabilitation Sciences (NIH T32HD057850), and the “Translating Obesity, Metabolic Dysfunction, and Comorbid Disease States” training program (NIH T32DK128770).

## Acknowledgments

ST and DEW designed the research study; all authors contributed to screening publications and data generation; ST analyzed the data and generated the figures, and all authors contributed to the manuscript.

## Data Availability Statement

The raw data supporting the conclusions of this article will be made available by the authors, without undue reservation.

## Notes

### Competing Interest Statement

The authors have declared no competing interest.

### Author Declarations

All papers included in our analysis were found on PubMed. A copy of our EndNote library is available upon request.

