## Supplementary Figures for "Abnormal Intraepidermal Nerve Fiber Density in Disease: A Scoping Review"

**Table 1:** Human Disease Classifications

| **Autoimmune** | **Neurodegenerative** | **Hereditary** |
| --- | --- | --- |
| Acute Inflammatory Demyelinating Polyradiculoneuropathy | Amyloid Neuropathy | CADASIIL |
| Anti–Myelin-Associated Glycoprotein (anti-MAG) Neuropathy | Amyotrophic Lateral Sclerosis | Charcot–Marie–Tooth Disease |
| Chronic Inflammatory Demyelinating Polyradiculoneuropathy | Eosinophilia-Associated Neuropathy | Epidermolysis Bullosa |
| Contactin-Associated Protein-Like 2 Autoantibodies | Ganglionopathy | Fabry Disease |
| Latent Autoimmune Diabetes of Adulthood | Multiple Systems Atrophy | Familial Amyloid Polyneuritis |
| Leucine-Rich Glioma Inactivated 1 Autoantibodies | Neuropathy of Unspecified Origin | Familial Dysautonomia |
| Lupus | Parkinson's Disease | Friedreich’s Ataxia |
| Miller Fisher Syndrome | POEMS Syndrome | Pachyonychia Congenita |
| Multiple Sclerosis | Supranuclear Palsy | Pompe Disease |
| Rheumatoid Arthritis | Paresthesia | Rett Syndrome |
| Sjögren's Syndrome | Polyneuropathy |  |
|  | Tauopathy |  |
| **Infectious** | **Metabolic** | **Drug Induced** |
| Human Immunodeficiency Virus | Diabetes Mellitus | Capsaicin Patch |
| Leprosy | Impaired Glucose Tolerance | Chemotherapy |
| Postherpetic Neuralgia | Metabolic Syndrome | Lidocaine Treatment |
|  | Obesity |  |
|  | Prediabetes |  |
| **Other** | **Other** | **Other** |
| >60 Years of Age | Heavy Alcohol Use | Prurigo |
| Atopic Dermatitis | Hypothyroidism | Pruritis |
| Autism | Idiopathic Chronic Itch | Psoriasis |
| Brachioradial Pruritus | Lichen Sclerosis | REM Sleep Disorder |
| Carpal Tunnel Syndrome | Light Chain Amyloidosis | Restless Leg Syndrome |
| Chronic Kidney Disease | Monopolar Depression | Sarcoidosis |
| Complex Regional Pain Syndrome | Non-Freezing Cold Injury | Sepsis |
| Cutaneous Amyloidosis | Occupational Exposure to Hand-Transmitted Tools | Transthyretin Amyloidosis |
| Endometriosis w/ Leg Pain | Peripheral Artery Disease | Whiplash Associated Disorder |
| Fibromyalgia | Postural Tachycardia Syndrome |  |

**Table 2:** Rodent Disease Classifications

| **Drug Induced** | **Metabolic** | **Infectious** |
| --- | --- | --- |
| Acrylamide Treatment | Ketogenic Diet | HIV-Tat Induction |
| Antibiotic | Obesity Model | Varicella Zoster Virus |
| Antiviral | Obesogenic Diet |  |
| Capsaicin Analog Treatment | Type 1 DM Model |  |
| Chemotherapy | Type 2 DM Model |  |
| MRI Contrast Agent |  |  |
| Resiniferatoxin Treatment |  |  |
| **Hereditary** | **Neurodegenerative** |  |
| Familial Amyloid Polyneuropathy Model | ALS Model |  |
| Recessive Dystrophic Epidermolysis Bullosa Model |  |  |
| **Other** | **Other** | **Other** |
| 100g Weight Traction Injury | Diphtheria Toxin Receptor Targeted to Langerhans Cells | Mercury Chloride Exposure |
| Chronic Constriction Injury | Dry Skin Model | Monoiodoacetate Treatment |
| Laser Induced Neuropathy | Electrical Shock | NaV1.7 protein KO |
| Nerve Injury | Femoral Artery Ligation | NAV1.8 Channel Mutation |
| Acetone Dry Skin Model | Glucagon Gene-Derived Peptides Deficient Mice | Nerve Growth Factor Administration |
| Antigen Indiced Arthritis | Hypoxia-Inducible Factor 1 KO in peripheral sensory neurons | Psoriasis Model |
| Dermatitis Model | Insular Glutamate Transport Inhibitor Injection | Schwann Cell NMDA-R-Deficient Mice |
| Complement Freund’s Adjuvant | Insulin Receptor KO |  |
| Dermatitis Model | MAPK/ERK activation in Schwann Cells |  |
